# TCA cycle remodeling drives proinflammatory signaling in humans with pulmonary tuberculosis

**DOI:** 10.1101/2021.01.23.21250380

**Authors:** Jeffrey M. Collins, Dean P. Jones, Ashish Sharma, Manoj Khadka, Ken Liu, Russell R. Kempker, Brendan Prideaux, Kristal Maner-Smith, Nestani Tukvadze, N. Sarita Shah, James C.M. Brust, Rafick P. Sekaly, Neel R. Gandhi, Henry M. Blumberg, Eric Ortlund, Thomas R. Ziegler

**Author notes:** Co-senior authors (Eric Ortlund, Thomas R. Ziegler).

## Abstract

The metabolic signaling pathways that drive pathologic tissue inflammation and damage in humans with pulmonary tuberculosis (TB) are not well understood. Using combined methods in plasma high-resolution metabolomics, lipidomics and cytokine profiling from a multicohort study of humans with pulmonary TB disease, we discovered that IL-1β-mediated inflammatory signaling was closely associated with TCA cycle remodeling, characterized by accumulation of the pro-inflammatory metabolite succinate and decreased concentrations of the anti-inflammatory metabolite itaconate. This inflammatory metabolic response was particularly active in persons with multidrug-resistant (MDR)-TB that received 2 months of ineffective treatment and was only reversed after 1 year of appropriate anti-TB chemotherapy. Both succinate and IL-1β were significantly associated with proinflammatory lipid signaling, including increases in the products of phospholipase A2, increased arachidonic acid formation, and metabolism of arachidonic acid to proinflammatory eicosanoids. Together, these results indicate that decreased itaconate and accumulation of succinate and other TCA cycle intermediates are important drivers of IL-1β-mediated proinflammatory eicosanoid signaling in humans with pulmonary TB disease. Host-directed therapies that mitigate such metabolic reprograming have potential to limit pulmonary inflammation and tissue damage.

**Graphical Abstract:** 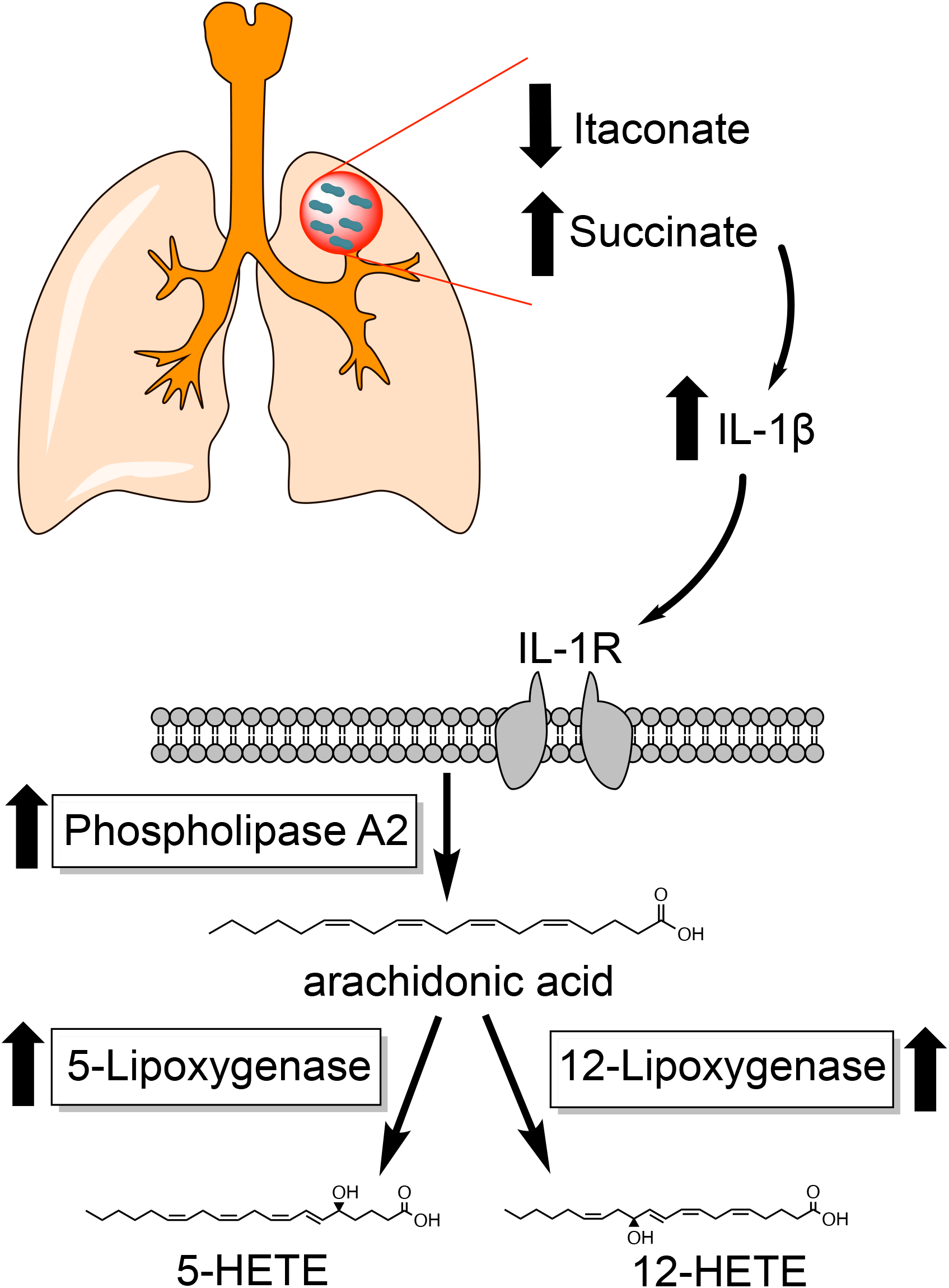

**One Sentence Summary:** Remodeling of the tricarboxylic acid cycle, characterized by increases in the proinflammatory metabolite succinate and decreased itaconate, mediates proinflammatory eicosanoids signaling in humans with pulmonary tuberculosis through induction of IL-1β.

## INTRODUCTION

Prior to the COVID-19 pandemic, tuberculosis (TB) was the leading global cause of infectious disease mortality, accounting for ∼1.4 million deaths each year (*1*). In persons successfully treated for pulmonary TB, rates of post-TB obstructive lung disease are high (*2, 3*) and may be even higher in persons treated for multidrug resistant (MDR)-TB (*4*). Such long-term changes in pulmonary function are likely the result of pathologic lung inflammation and tissue damage prior to and shortly after initiation of anti-TB chemotherapy (*5*). Elucidating host response pathways that promote pathologic tissue inflammation in persons with pulmonary TB will be critical to detect those at greatest risk for adverse pulmonary outcomes and identify targets for host-directed therapeutics.

The cytokine IL-1 plays a complex role following infection with *Mycobacterium tuberculosis* (*Mtb*) that appears to evolve as one progresses to TB disease. In mice infected with *Mtb*, IL-1 initially augments production of anti-inflammatory eicosanoid prostaglandin E2 (PGE2) and limits production of type I interferons thereby enhancing control of bacterial replication (*6*). However, in later stages of TB disease, IL-1 signaling leads to upregulation of proinflammatory eicosanoids and an influx of neutrophils, which promote further bacterial replication and tissue destruction (*7*). Similarly, IL-1 signaling in macaques with TB disease is highly correlated with pulmonary inflammation and IL-1 blockade attenuates tissue damage (*8*). This suggests that as TB disease progresses, IL-1 signaling becomes pathologic and leads to tissue inflammation that is permissive to bacterial growth. The primary drivers of IL-1 signaling in humans with pulmonary TB and whether such signaling yields a primarily proinflammatory or anti-inflammatory phenotype has yet to be established.

Recent studies in macrophage biology show tricarboxylic acid (TCA) cycle remodeling may play an important role in regulating IL-1 (*9-11*). Inflammatory macrophage activation leads to accumulation of TCA cycle intermediates such as succinate, which, in turn, results in upregulation of the proinflammatory IL-1β-HIF-1∝axis (*9-11*). This metabolic remodeling is regulated by the metabolite itaconate, which inhibits succinate dehydrogenase and stabilizes the anti-inflammatory transcription factor Nrf2, thereby limiting the proinflammatory cascade (*9, 12*). However, the contribution of this host metabolic response pathway to inflammation in human pulmonary TB has not been previously described.

To better characterize the contribution of host metabolism to pathologic inflammatory cascades in humans with pulmonary TB, we performed detailed immunometabolic profiling using combined approaches in targeted and untargeted high-resolution metabolomics (HRM), lipidomics and cytokine profiling in a multicohort study of persons with drug susceptible (DS) and MDR pulmonary TB. Our aim was to gain a more comprehensive understanding of the metabolic drivers of inflammation in TB disease and elucidate new targets for host-directed therapeutics to limit inflammation and tissue damage. Here we show TCA cycle remodeling, characterized by accumulation of TCA cycle intermediates and decreased itaconate, is closely associated with molecular response networks of pathologic inflammation in human TB disease.

## RESULTS

### Metabolic Pathway Regulation in MDR-TB

We hypothesized that persons experiencing delays in the receipt of effective anti-TB chemotherapy would experience continued progression of pulmonary TB disease and propagation of proinflammatory signaling cascades. We therefore sought to determine which metabolic pathways were most regulated in a population with MDR-TB (n=85) that was initially treated with 2 months of ineffective, first-line anti-TB drug therapy prior to the diagnosis of MDR-TB (**Table 1**; described in Methods) (*13*). Persons with MDR-TB were enrolled from 2011-2013 and prior to widespread implementation of rapid molecular diagnostic testing (Xpert MTB/RIF). The diagnosis of MDR-TB was made after a positive sputum culture for *Mtb* demonstrated MDR-TB using phenotypic drug susceptibility testing (DST). We compared the plasma metabolome in the MDR-TB cohort to a population of persons with drug susceptible (DS)-TB (n=89) enrolled within 1 week of pulmonary TB diagnosis as well as a group of control participants (n = 57), which included asymptomatic individuals with LTBI (n = 20) and those without evidence of TB disease or *Mtb* infection (n = 37) (*14*).

**Table 1.** Clinical and demographic characteristics of study participants.

| **Participant Characteristics** | **MDR-TB cohort**<br>**KwaZulu-Natal,**<br>**South Africa (n=85)** | **DS-TB cohort**<br>**Tbilisi, Georgia**<br>**(n=89)** | **Controls without**<br>**active TB disease**<br>**(n=57)** |
| --- | --- | --- | --- |
| Female sex, n (%) | 53 (62) | 31 (35) | 40 (70) |
| Age, years (median [IQR]) | 34 (27-42) | 31 (24-43) | 46 (37-54) |
| HIV positive, n (%) | 64 (75) | 0 (0) | 1 (2) |
| AFB sputum smear positive at diagnosis, n (%) | N/A | 89 (100) | N/A |
| AFB sputum smear positive at first study visit, n (%) | 30 (45.5)* | 67 (75) | N/A |
| Sputum culture positive for *M. tuberculosis* at diagnosis, n (%) | 85 (100) | 89 (100) | N/A |
| Sputum culture positive for *M. tuberculosis* at first study visit, n (%) | 56 (78.9)** | 76 (85) | N/A |
| Time to sputum culture conversion, days (median [IQR]) | 62 (51-87)§ | 27.5 (21.5-46.5) | N/A |
| Positive test for latent TB infection, n (%) | N/A | N/A | 20 (35.1) |

Multidrug resistant (MDR); Drug susceptible (DS)
\*AFB sputum smear results were not available for 19 participants (22.4%)
\*\*AFB sputum culture results were not available for 14 participants (16.5%)
§Data on sputum culture conversion was missing for 7 participants (12.5%) with a positive sputum culture at baseline

With plasma HRM, we detected 9,787 m/z features using C18 liquid chromatography in negative ionization mode, of which 5,146 were present in >90% of samples and selected for downstream analysis. We first sought to determine which metabolic pathways were significantly regulated in persons with MDR-TB versus controls and persons with DS-TB. After adjustment for age, sex and HIV status, unbiased pathway analysis (*15*) revealed the TCA cycle as well as multiple overlapping pathways in fatty acid metabolism including de novo fatty acid biosynthesis, fatty acid activation, and fatty acid oxidation, were the most significantly regulated metabolic pathways in persons with MDR-TB versus controls without TB disease (**Fig. 1A**) as well as persons with DS-TB (**Fig. 1B**). We then performed a correlation analysis of metabolites with primary adducts (M-H) mapping to significant metabolic pathways. This analysis yielded a cluster of highly correlated metabolites that were upregulated in the MDR-TB cohort including arachidonic acid, homolinoleic acid and TCA cycle intermediates alpha-ketoglutarate, succinate, fumarate and malate (p<0.001 for all; **Fig. 1C**). There was a smaller cluster of correlated metabolites mapping to the xenobiotics and tyrosine metabolic pathways that were down-regulated in MDR-TB. Because of the high prevalence of HIV co-infection in the MDR-TB cohort, we performed a principal component analysis of identified metabolites to ensure that observed differences were not driven by HIV status. We found persons with MDR-TB with and without HIV co-infection clustered together and both groups clustered separately from persons with DS-TB and controls, suggesting the observed differences were not related to HIV status (**Fig. S1**).

**Fig. 1.**
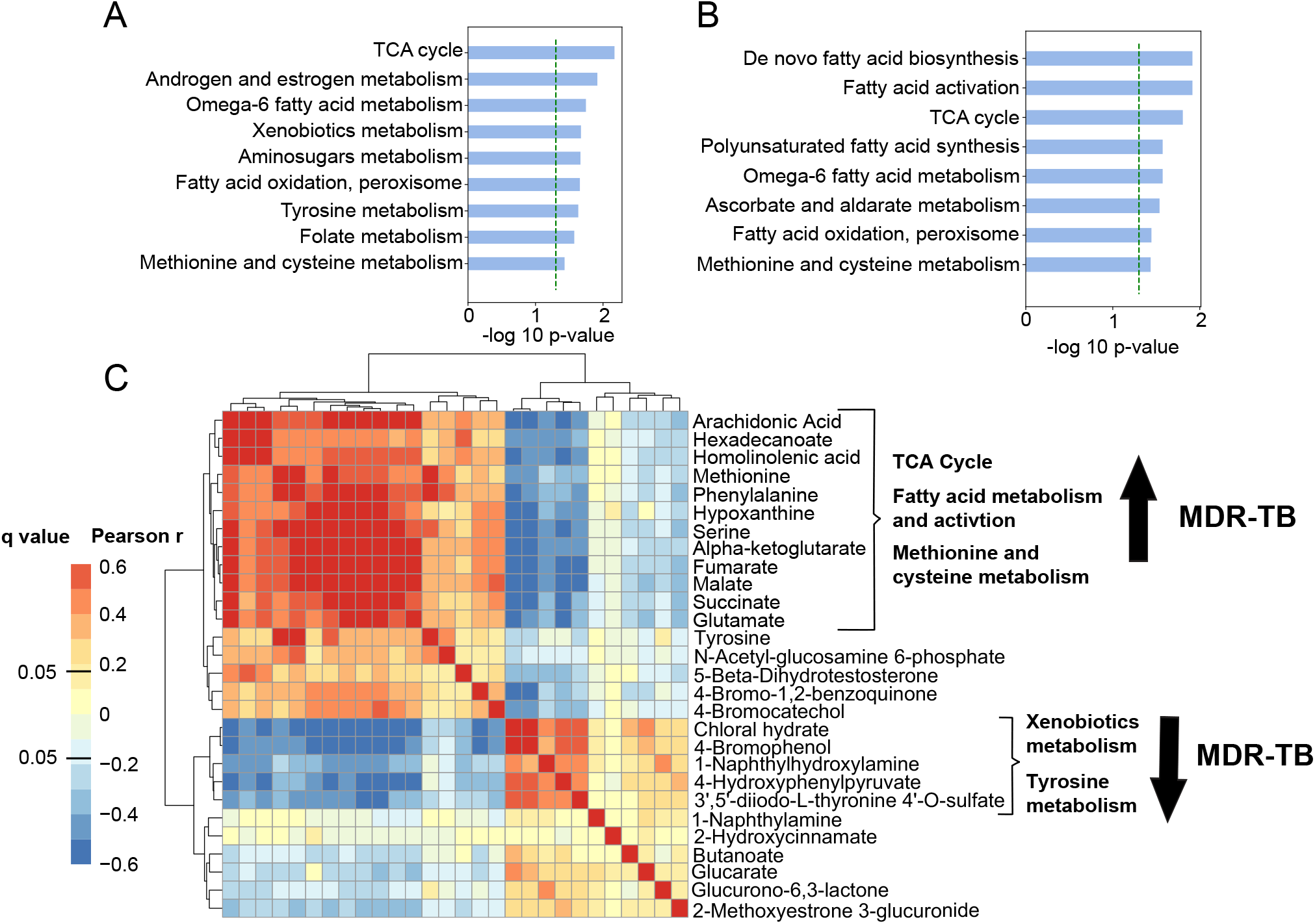
Metabolic pathway regulation in MDR-TB. (A) Unbiased pathway analysis using plasma HRM with C18 negative chromatography to compare the metabolome of persons with multidrug resistant (MDR)-TB (n=85) to (A) asymptomatic controls without TB disease (n=57) and (B) persons with drug susceptible (DS)-TB (n=89) shows TCA cycle metabolism and multiple overlapping pathways in fatty acid synthesis, activation and metabolism were the most significantly regulated pathways in MDR-TB. The y-axis shows significantly regulated metabolic pathways and the x-axis shows the -log p-value for pathway enrichment adjusted for age, sex and HIV status. (C) Correlation analysis of primary metabolite adducts (M-H) mapping to significant pathways reveals a network of highly correlated metabolites involved in the TCA cycle, fatty acid metabolism and methionine and cysteine metabolism that were significantly increased in persons with MDR-TB.

Accumulation of succinate and downstream TCA cycle intermediates has been described as a potent driver of inflammation in multiple disease states (*9, 10, 16*). We therefore hypothesized that TCA cycle remodeling in MDR-TB represented a pro-inflammatory signaling cascade. To better characterize observed changes in this metabolic pathway, TCA cycle intermediates were confirmed and quantified by accurate mass, MS/MS and retention time relative to authentic standards (*17*). TCA cycle remodeling in MDR-TB was characterized by significant increases in plasma concentrations of succinate, fumarate and malate versus persons with DS-TB and asymptomatic controls (p<0.001 for all; **Fig. 2**). Persons with MDR-TB also exhibited significant increases in plasma concentrations of alpha-ketoglutarate and glutamate (p<0.001 for all) whereas concentrations of citrate were significantly decreased in persons with MDR-TB and concentrations of cis-aconitate were similar between groups. These findings suggest the primary source of succinate in those with MDR-TB was not flux through the TCA cycle, but rather glutamine-dependent anaplerosis to alpha-ketoglutarate as has been described in *in vitro* and mouse models (*10*).

**Fig. 2.**
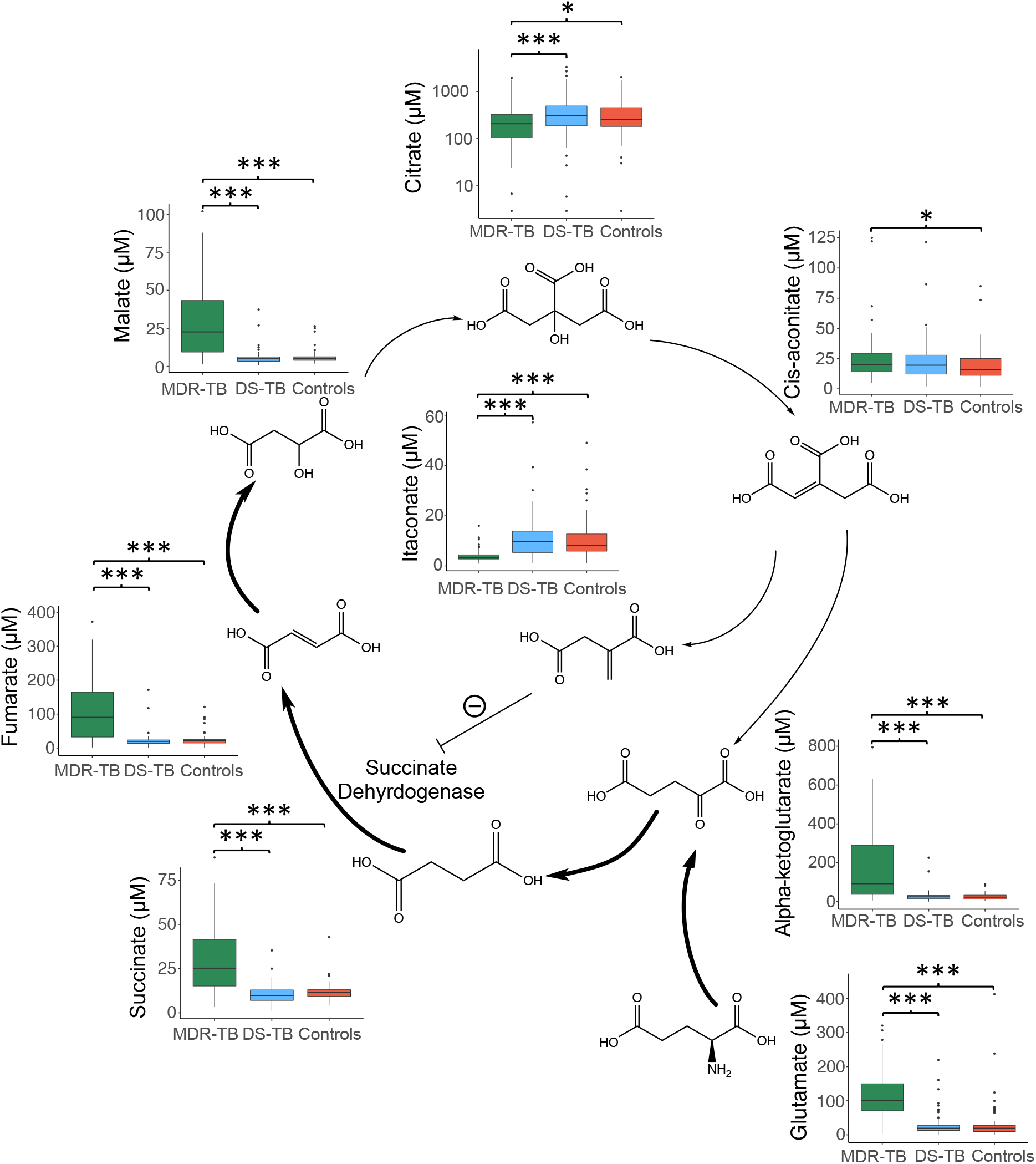
Characterization of TCA cycle remodeling in MDR-TB. Plasma quantification of TCA cycle metabolites in persons with multidrug resistant (MDR)-TB shows significant increases in the metabolites alpha-ketoglutarate, succinate, fumarate and malate versus persons with drug susceptible (DS)-TB and controls without TB disease. Plasma concentrations of glutamate were also significantly increased in MDR-TB while citrate was significantly decreased and concentrations of cis-aconitate were similar between groups. Plasma concentrations of the anti-inflammatory metabolite itaconate were significantly decreased in persons with MDR-TB. Groups were compared using the Wilcox rank sum test: * p<0.05, ** p<0.01, *** p<0.001.

The metabolite itaconate is derived from TCA cycle intermediate cis-aconitate and has been shown to attenuate inflammatory metabolic signals through inhibition of succinate dehydrogenase (*9*) and induction of Nrf2 (*12*). Thus, we posited that accumulation of TCA cycle intermediates in MDR-TB may be due, in part, to decreases in itaconate. Indeed, we found itaconate was significantly decreased in persons with MDR-TB compared to persons with DS-TB and controls (p<0.001). Plasma concentrations of itaconate were significantly and negatively correlated with plasma concentrations of succinate, fumarate and malate (p<0.001 for all; data not shown) consistent with a role in negatively regulating these metabolites. These findings suggest persons with progressive and untreated pulmonary TB experience proinflammatory metabolic remodeling characterized by accumulation of succinate via glutamine-dependent anaplerosis and decreased itaconate.

### TCA cycle remodeling is associated with levels of IL-1β

Accumulation of TCA cycle intermediates such as succinate drive inflammation by increasing production of IL-1β (*9, 10, 16*). We therefore hypothesized that the primary role of TCA cycle remodeling seen in participants with MDR-TB was induction of IL-1β. To answer this question, we measured plasma cytokine concentrations in a subset of persons with MDR-TB (n=37), persons with DS-TB (n=29) and persons without TB disease (n=20). We found concentrations of IL-1β were highly correlated with plasma concentrations of TCA cycle intermediates and negatively correlated with itaconate (p<0.001 for all; **Fig. 3A**). Using hierarchical clustering analysis (HCA) of correlations between plasma cytokines and TCA cycle intermediates, we found IL-8 and IL-4 were also highly correlated with TCA cycle intermediates and formed a cluster with IL-1β. Plasma concentrations of IL-1β were significantly elevated in both persons with MDR-TB and DS-TB versus controls (p<0.001 and p=0.03 respectively), though the elevation was significantly greater in persons with MDR-TB (p<0.001; **Fig. 3B**). Plasma concentrations of IL-1β were highly correlated with the plasma ratio of succinate to itaconate (p<0.001; **Fig. 3C**). For those participants with available culture conversion data, plasma concentrations of IL-1β as well as plasma concentrations of the succinate/itaconate ratio at study enrollment were significantly and positively correlated with subsequent time to sputum culture conversion (p<0.001 for both; **Fig. 3D** and **3E**). Additionally, persons with DS-TB and a persistently positive AFB sputum smear at study enrollment (within 1 week of treatment start) had a significantly higher plasma succinate/itaconate ratio versus those who had already converted to a negative AFB sputum smear (p=0.04; **Fig. S2)**. These findings indicate that TCA cycle remodeling in pulmonary TB drives inflammation through production of IL-1β and is associated with delayed treatment response.

**Fig. 3.**
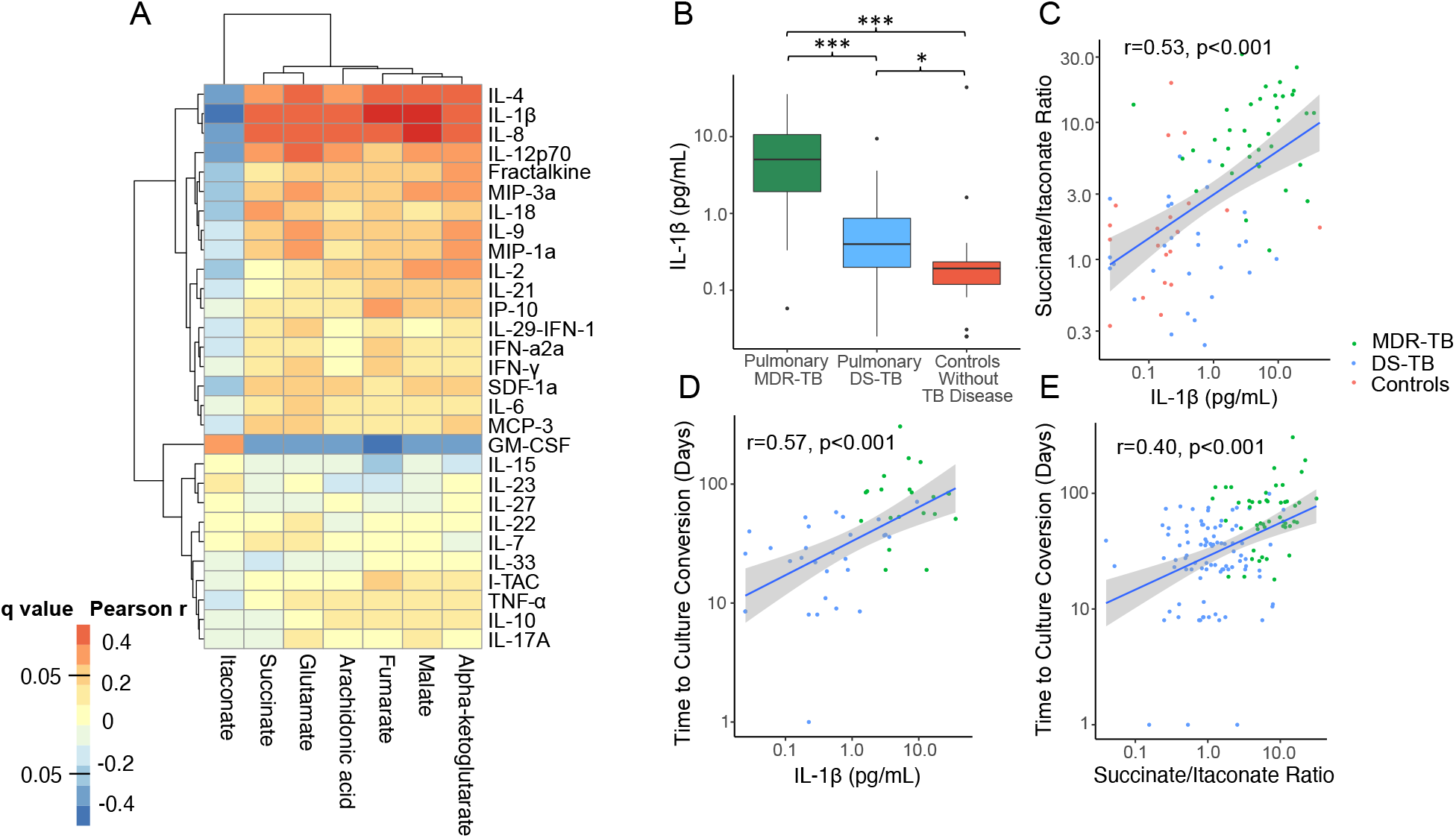
TCA cycle remodeling is associated with increased IL-1β and delayed response to treatment. (A) In a subset of persons with MDR-TB (n=37), DS-TB (n=29) and controls without TB disease (n=20), plasma concentrations of TCA cycle intermediates and arachidonic acid were highly correlated with concentrations of IL-1β, IL-4 and IL-8. (B) Plasma concentrations of IL-1β were significantly higher in person with MDR-TB versus those with DS-TB and controls. Plasma concentrations of IL-1β were (C) significantly and positively correlated with the plasma ratio of succinate/itaconate. Plasma concentrations of (D) IL-1β as well as (E) the plasma succinate/itaconate ratio at enrolment were significantly and positively correlated with time to sputum culture conversion. Groups were compared using the Wilcox rank sum test: * p<0.05, ** p<0.01, *** p<0.001.

### TCA cycle remodeling drives proinflammatory lipid signaling through IL-1β

In early *Mtb* infection, IL-1β shifts the balance of eicosanoid production toward anti-inflammatory prostaglandin E2 and away from proinflammatory eicosanoids, thereby limiting *Mtb* replication (*6*). However, in later stages of TB disease, IL-1β signaling is associated with eicosanoid-mediated inflammation and tissue damage (*7, 8, 18*). Given plasma concentrations of IL-1β and TCA cycle intermediates were highly correlated with concentrations of arachidonic acid, we posited that TCA cycle remodeling and IL-1β were acting primarily to drive proinflammatory lipid signaling cascades in pulmonary TB disease in humans. To examine this question, we first sought to determine whether persons with MDR-TB exhibited increased phospholipase A2 activity, the first step in formation of arachidonic acid (AA). We performed untargeted lipidomics on plasma samples from a subset of the study population including persons with MDR-TB (n=50), persons with DS-TB (n=30) and controls without TB disease (n=20). Of plasma lipids with confirmed chemical identities, persons with MDR-TB demonstrated significant upregulation of multiple species of lyso-phospholipids and significant decreases in phospholipids with two acyl chains compared to persons without TB disease (**Fig. 4A)** and persons with DS-TB (**Fig. 4B**), consistent with significant increases in phospholipase A2 activity.

**Fig. 4.**
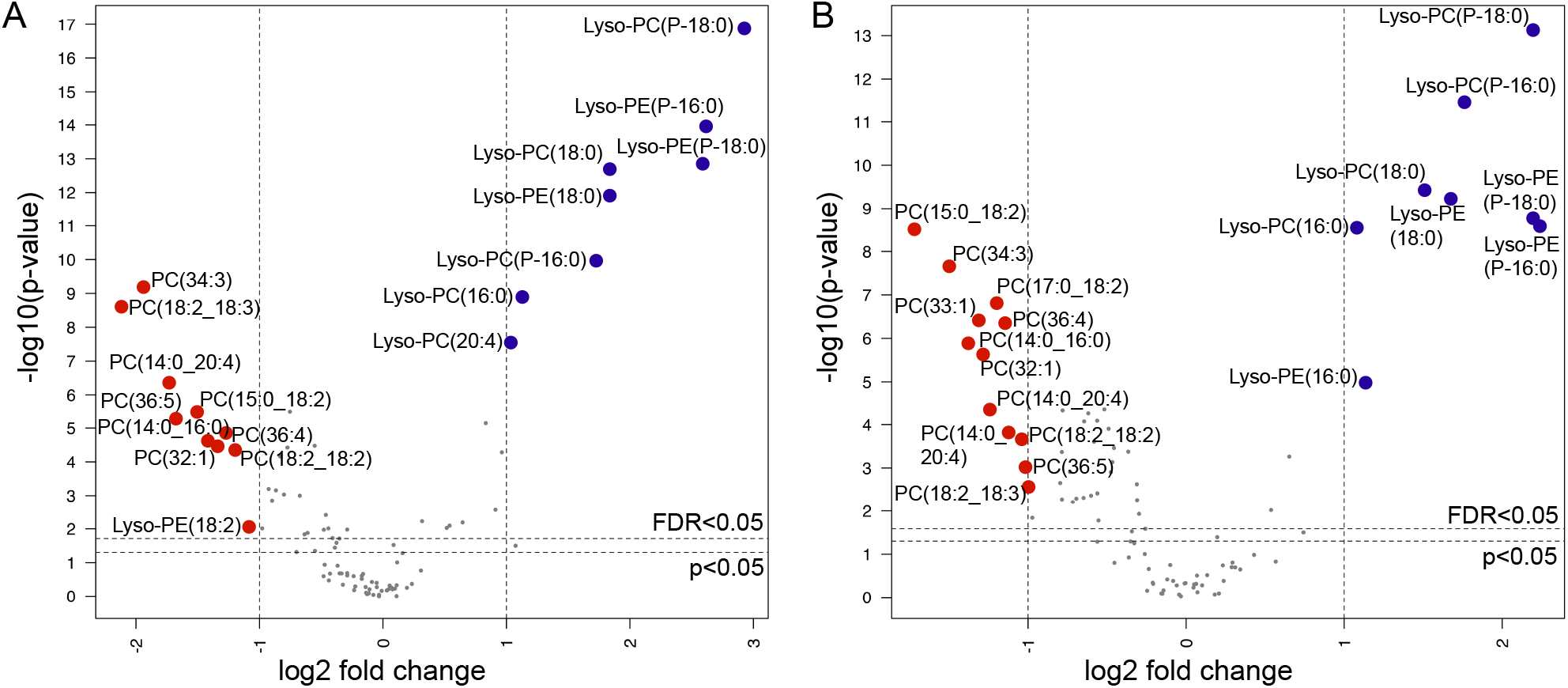
Increased phospholipase activity in MDR-TB. Volcano plots of plasma lipids with confirmed chemical identities shows significant increases of lyso-phospholipids and significant decreases in phospholipids with two acyl chains in persons with MDR-TB (n=50) versus (A) asymptomatic controls without TB disease (n=20) and (B) persons with drug susceptible (DS)-TB (n=30). The x-axis shows the log2 fold change in expression in MDR-TB versus controls and DS-TB for each lipid and the y-axis shows the -log p-value for each comparison after adjustment for age, sex and HIV status. Lipids upregulated in MDR-TB with >1 log2 fold change at a false discovery rate (FDR) (*19*) of q<0.05 are labelled in blue and those downregulated are labelled in red.

Following formation from phospholipase A2, AA is further metabolized through one of three metabolic pathways to regulate inflammation: cyclooxygenases (COX) to form anti-inflammatory prostaglandins, lipoxygenases (LOX) to form proinflammatory eicosanoids and CYP450 enzymes to form the less biologically active dihydroxyeicosatrienoic acids (DHETs) (*20*). Prior studies have shown that proinflammatory eicosanoids formed through the LOX pathway are significantly upregulated in persons with pulmonary TB disease and associated with more severe clinical disease (*6, 7, 21*) and cavity formation (*22*). We therefore hypothesized that accumulation of TCA cycle intermediates, IL-1β and arachidonic acid in MDR-TB patients were part of an inflammatory cascade resulting in increased production of proinflammatory eicosanoids. To further characterize downstream changes in AA metabolism in persons with MDR-TB versus those with DS-TB and asymptomatic controls, we performed a targeted oxylipin assay to identify and quantify relevant eicosanoids in plasma samples (*23*). All persons with pulmonary TB demonstrated significant increases in 11,12-DHET and 14,15-DHET versus controls (p<0.001 for both; **Fig. 5A** and **5B**), consistent with increases in AA metabolism. However, while concentrations of the less biologically active DHET molecules did not differ between TB disease groups, persons with MDR-TB demonstrated significant increases in the proinflammatory eicosanoids 5-HETE and 12-HETE compared to both persons with DS-TB and controls (p<0.001 for all; **Fig. 5C** and **5D**). This indicates AA metabolism in persons with MDR-TB was both increased and more likely to produce proinflammatory eicosanoids metabolized via LOX pathways compared to persons with DS-TB. Plasma concentrations of 5-HETE and 12-HETE, but not 11,12-DHETE or 14,15-DHETE, were highly correlated with plasma concentrations of succinate, fumarate and malate as well as IL-1β, IL-8 and IL-4 while they were negatively correlated with concentrations of itaconate (p<0.001 for all; **Fig. 5E**). These findings suggest TCA cycle remodeling and IL-1β drive proinflammatory lipid signaling in pulmonary TB.

**Fig. 5.**
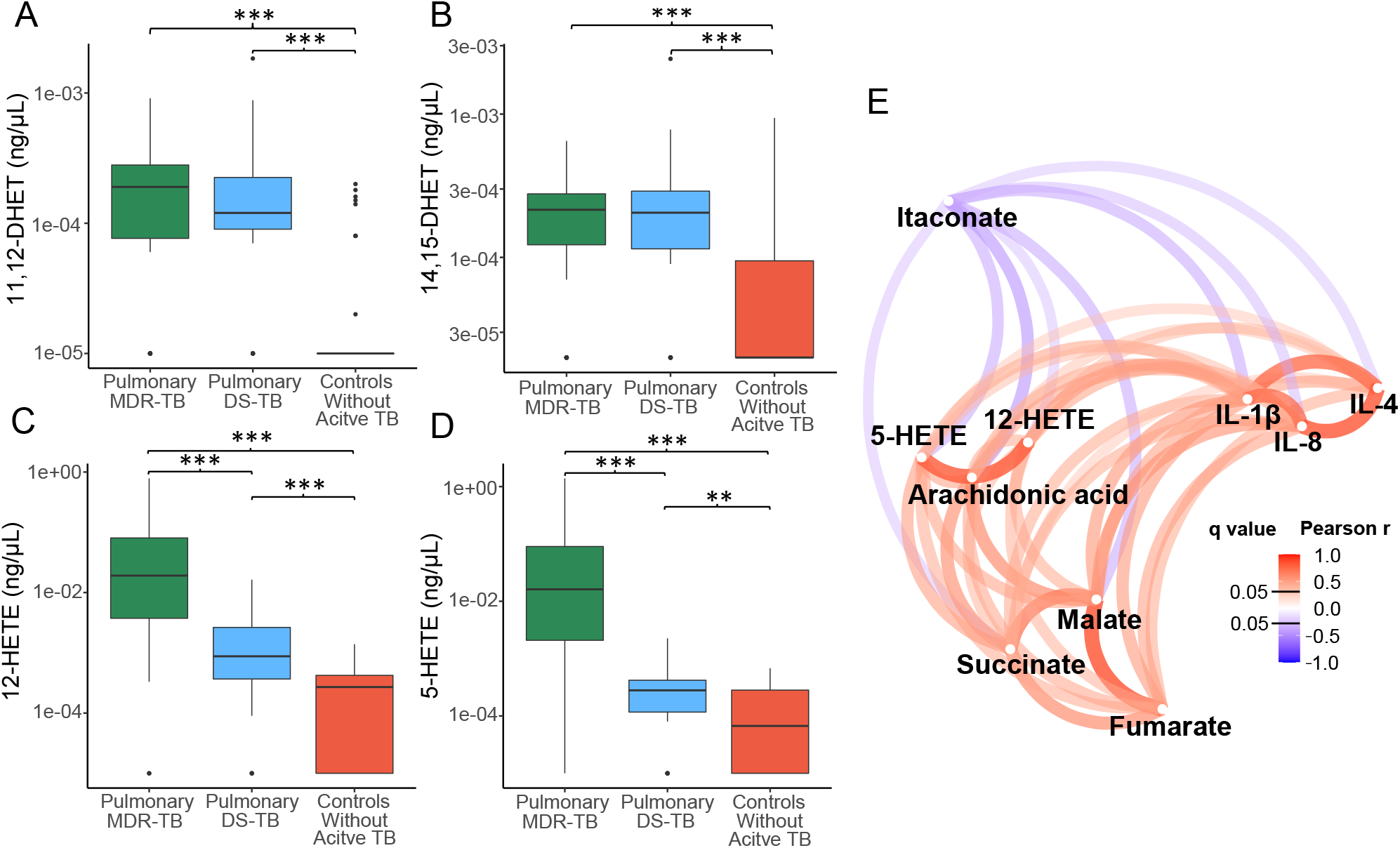
Immonometabolic remodeling is associated with proinflammatory eicosanoid signaling. Compared to asymptomatic controls without TB disease (red; n=39), persons with multidrug resistant (MDR)-TB (green; n=50) as well as persons with drug susceptible (DS)-TB (blue; n=30) demonstrated significant increases in plasma concentrations of the arachidonic acid (AA) metabolites (A) 11,12-DHET and (B) 14,15-DHET metabolized by the CYP450 system. Proinflammatory eicosanoids including (C) 5-HETE and (D) 12-HETE produced by lipoxygenase metabolism of AA were significantly increased in persons with MDR-TB versus persons with DS-TB and asymptomatic controls. (E) Shows a network plot of metabolites and cytokines correlated with plasma concentrations of 5-HETE and 12-HETE with dark red indicating a higher Pearson correlation coefficient and dark blue indicating a more negative correlation. Plasma concentrations of 5-HETE and 12-HETE had strong positive correlations with plasma concentrations of arachidonic acid, succinate, fumarate, malate, IL-1β, IL-8 and IL-4. Conversely, plasma concentrations of 5-HETE and 12-HETE were negatively correlated with plasma concentrations of itaconate. Groups were compared using the Wilcox rank sum test: * p<0.05, ** p<0.01, *** p<0.001.

### Proinflammatory metabolic remodeling is reversed with TB treatment

Because patients with MDR-TB were enrolled from a different country from those with DS-TB and controls without TB disease, we examined the possibility that observed differences in this population could be due to factors unrelated to TB disease. We reasoned that if TCA cycle remodeling were due to TB disease then these changes would be reversed with appropriate anti-TB chemotherapy. We therefore examined the change in plasma concentrations of metabolites and cytokines in a subset of MDR-TB patients where plasma samples were available over the first year of treatment (n=17).

While there was minimal change in plasma concentrations of succinate at 2-4 months, there was a significant decline after 1 year of appropriate treatment for MDR-TB (p=0.02; **Fig 6A**). There was an increase in plasma itaconate concentrations after 2-4 months of treatment and 1 year of treatment that did not meet statistical significance (p=0.06 and 0.12 respectively; **Fig 6B**). Similar to succinate, plasma concentrations of arachidonic acid and IL-1β significantly declined relative to baseline after 1 year of MDR-TB treatment (p=0.03 and p=0.003 respectively; **Fig 6C** and **6D**). Using principal component analysis of the inflammatory network including TCA cycle metabolites, IL-1β, IL-4, IL-8 and arachidonic acid, we found the MDR-TB group clustered separately from those with DS-TB and controls at baseline, but clustered more closely with these groups after 1 year of MDR-TB treatment. (**Fig 6E**). Together, these findings indicate that proinflammatory metabolic remodeling is reversed with appropriate TB treatment, but that the timeline of this reversal may be prolonged.

**Fig. 6.**
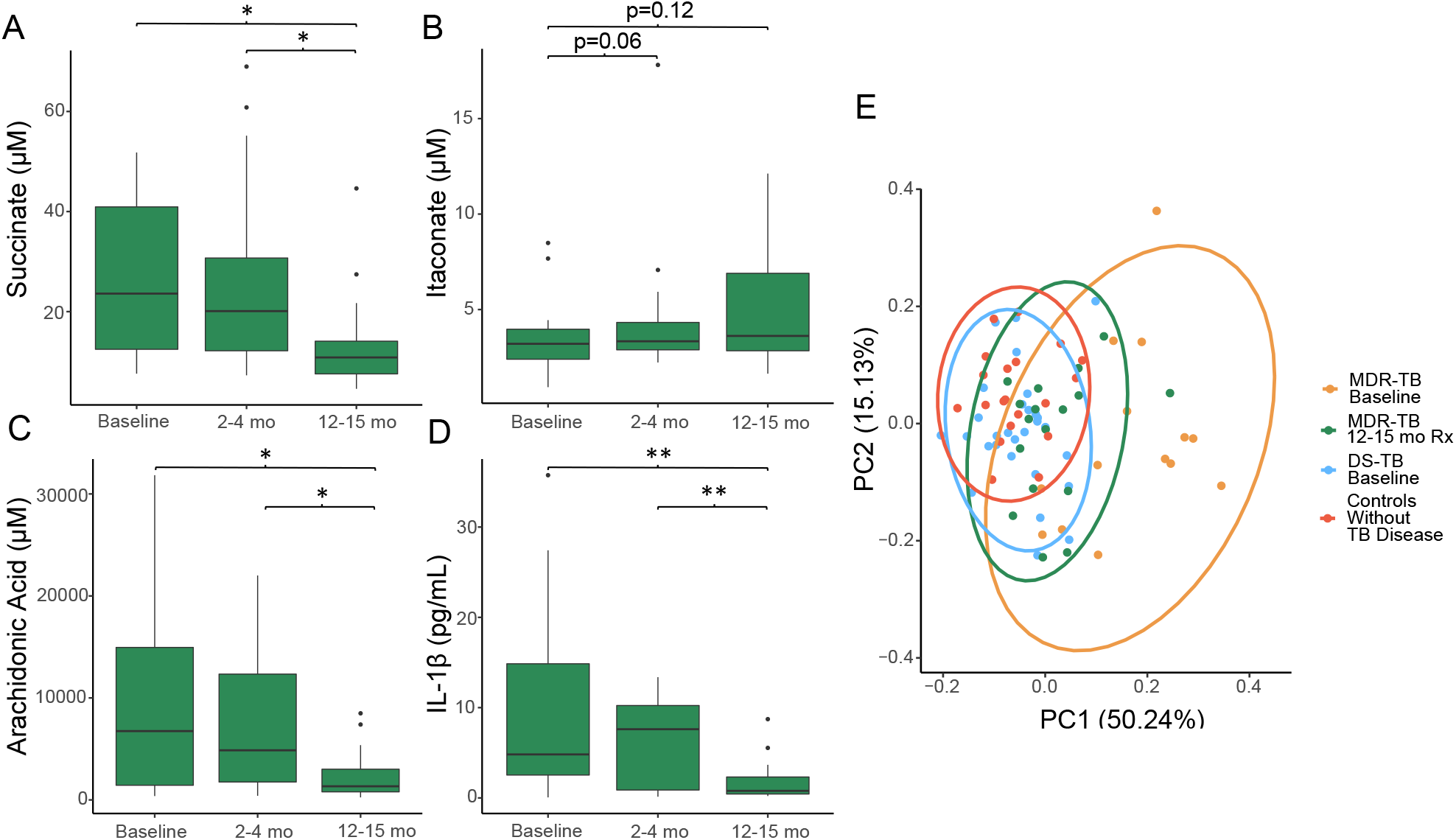
Proinflammatory immunometabolic changes are reversed with appropriate TB treatment. (A) In persons with multidrug resistant (MDR)-TB (n=17) here was a significant decline in plasma succinate concentration after 1 year of TB therapy while (B) there was an increase in plasma concentrations of itaconate after 2-4 months and 1 year of treatment that did not meet statistical significance. Plasma concentrations of (C) arachidonic acid (D) IL-1β also significantly declined after 1 year of treatment. (E) Using principal component analysis of TCA cycle metabolites, arachidonic acid and IL-1β, IL-4 and IL-8, we found after 1 year treatment the MDR-TB group clustered with controls and those with DS-TB. Groups were compared using the Wilcox signed rank test: * p<0.05, ** p<0.01, *** p<0.001.

## DISCUSSION

In this multicohort study, we used an unbiased approach to plasma metabolic pathway analysis combined with targeted and untargeted lipidomics and cytokine profiling to discover that TCA cycle remodeling is strongly associated with IL-1β-mediated proinflammatory eicosanoid signaling in persons with pulmonary TB disease. We found this inflammatory cascade, characterized by increased plasma concentrations of TCA cycle intermediates succinate, fumarate and malate and decreased concentrations of itaconate, was particularly upregulated in persons with MDR-TB after 2 months of ineffective anti-TB chemotherapy and was reversed only after 1 year of efficacious treatment. Collectively, these results indicate that TCA cycle remodeling is an important driver of proinflammatory eicosanoid signaling in pulmonary TB disease. Furthermore, our findings suggest that prolonged delays in effective treatment allow this proinflammatory cascade to perpetuate, potentially contributing to poor treatment outcomes in MDR-TB. These findings also indicate that administration of host-directed therapies targeting TCA cycle remodeling, such as itaconate analogs (*12*), may be an effective way to reduce pathologic inflammation and limit tissue damage (*6-9*).

Accumulation of succinate is known to drive inflammation in macrophages through stabilization of the transcription factor HIF-1α to increase production of IL-1β (*10 16*). This metabolic remodeling is the result of a shift in cellular energy metabolism toward glycolysis and away from oxidative phosphorylation, thereby causing succinate and other TCA cycle intermediates to accumulate, a process termed aerobic glycolysis or the “Warburg Effect” (*24*). Prior studies have shown *Mtb* is able to induce aerobic glycolysis in pulmonary macrophages and that this change is necessary for production of IL-1β (*25 26*). The strong association between elevated succinate and IL-1β-mediated inflammatory signaling in our study suggests that TCA cycle remodeling plays a critical role in regulating the inflammatory response in humans with pulmonary TB. Further, our results indicate the primary source of TCA cycle intermediates in TB disease is glutamine-dependent anaplerosis, consistent with inflammatory metabolic remodeling driven by aerobic glycolysis (*10*).

However, while prior *in vitro* and mouse studies have shown induction of IL-1β results in control of *Mtb* replication (*6 25*), we found TCA cycle remodeling and induction of IL-1β produced a distinctly proinflammatory phenotype in humans that was associated with increased bacterial burden and prolonged time to sputum culture conversion. Our findings are supported by independent human studies, which have consistently show elevated plasma concentrations of IL-1β in persons with pulmonary TB are correlated with markers of inflammation (*27*) and associated with greater extent of disease and cavitation on chest radiograph (*18 28*). A possible explanation for these apparently discordant findings is that the role of IL-1β evolves following infection with *Mtb*. Initially, IL-1β provides a counterbalance to type-1-IFN signaling by promoting metabolism of AA to anti-inflammatory prostaglandins (*6*). Early IL-1 receptor blockade leads to greater abundance of proinflammatory eicosanoids and increased bacterial burden (*6 29*). In later stages of TB disease in mice, IL-1β becomes a driver of proinflammatory eicosanoid signaling leading to an influx of neutrophilic inflammation that is permissive to bacterial growth (*7*). In macaques with TB disease, plasma concentrations of IL-1β are highly correlated with levels of pulmonary inflammation and IL-1 receptor blockade limits tissue damage (*8*). Thus, accumulating TCA cycle intermediates and decreased itaconate during later stages of TB disease may be another example of *Mtb* exploiting a host metabolic adaption aimed at controlling *Mtb* replication during the initial stages of infection (*30 31*). While TCA cycle remodeling and increased IL-1β initially promotes control of bacterial replication (*25*), our findings suggest that as humans progress to TB disease it increases AA metabolism and conversion to proinflammatory eicosanoids, potentially worsening tissue damage and increasing disease severity.

This immunometabolic network has multiple potential targets where host-directed therapies could be used to limit pathologic inflammation. Similar to macaques with TB disease (*8*), our findings suggest IL-1 receptor blockade has potential to be therapeutic in humans with pulmonary TB by limiting inflammation and tissue damage. However, the large amount of heterogeneity we observed in IL-1β signaling between populations suggests this therapy may only benefit persons with more advanced disease or prolonged delays in treatment initiation. Our findings also indicate host-directed therapies directly targeting TCA cycle remodeling that are up-stream of IL-1β are also in need of further evaluation.Inhibition of succinate dehydrogenase, which regulates succinate levels (*9, 11, 24*), is one potential target that may limit host inflammation. Itaconate analogs such as 4-octyl itaconate (*12*) may also have potential to limit pulmonary inflammation by counteracting inflammatory signals driven by TCA cycle remodeling.

The observation that dysregulated IL-1β signaling occurs disproportionately in persons with MDR-TB has been reported previously (*27*). We hypothesize that in the present study this was caused by delays in adequate MDR-TB treatment, which allowed for propagation of a proinflammatory cascade. However, rifampin resistance mutations in *Mtb* isolates have been associated with differential metabolic responses and secretion of IL-1β in human macrophages (*32*). Thus, the possibility that the different host responses observed are related to differential induction of proinflammatory cascades by the organism itself cannot be ruled out.

This study is subject to several limitations. Though we demonstrate a strong association between plasma concentrations of TCA cycle intermediates, IL-1β and proinflammatory eicosanoids, the observational nature of the study precludes us from definitively establishing a causal relationship. While persons with MDR-TB were enrolled from three study sites, all were from the same country, so it remains possible that metabolic differences in this group are driven by host genetic differences or differences in environmental exposures rather than MDR-TB status. In future studies it will also be important to evaluate the contribution of the host microbiome, which accounts for a large proportion of succinate circulating in plasma (*33*).

In summary, we demonstrate that TCA cycle remodeling is closely associated with IL-1β-mediated proinflammatory eicosanoid signaling in humans with pulmonary TB. This remodeling is characterized by significant increases in plasma succinate concentrations and significant decreases in concentrations of itaconate. These findings provide evidence that pathologic eicosanoid signaling in pulmonary TB is driven by accumulation of TCA cycle intermediates. The TCA cycle may therefore represent a promising target for host-directed therapies aimed at limiting pulmonary inflammation and tissue damage.

## MATERIALS AND METHODS

### Sample collection

For all cohorts, blood was collected in ethylenediaminetetraacetic acid (EDTA)-containing tubes and centrifuged; isolated plasma was immediately frozen and stored at −80°C. Samples collected outside of the U.S. were subsequently shipped on dry ice to Emory University, Atlanta, GA, USA. All samples remained frozen during transit and were kept at −80°C prior to metabolomics, lipidomics and cytokine analysis.

### Multidrug resistant-TB cohort

Persons with pulmonary TB from KwaZulu-Natal province, South Africa were enrolled as part of a prospective, observational cohort study of MDR-TB and HIV co-infection from 2011-2013 (*13*). All persons in the South African cohort had MDR-TB as demonstrated by a positive sputum culture for *Mtb* and phenotypic DST indicating resistance to at least both isoniazid and rifampin. Baseline plasma samples were collected within 7 days of starting conventional treatment for MDR-TB. Persons with HIV-co-infection were continued on conventional anti-retroviral therapy (ART) and those not previously on ART were started on treatment. All patients were referred to a dedicated MDR-TB treatment center and treated with a standardized drug regimen that included kanamycin (15 mg/kg, maximum 1 g daily), moxifloxacin (400 mg daily), ethionamide (15– 20 mg/kg, maximum 750 mg daily), terizidone (15–20 mg/ kg, maximum 750 mg daily), ethambutol (15–20 mg/kg, maximum 1200 mg daily), and pyrazinamide (20–30 mg/kg, maximum 1600 mg daily). All persons who completed the study were treated for a period of two years and serial plasma samples were analyzed at enrollment and 2-4 months and 12-15 months after treatment initiation.

### Drug susceptible-TB cohort

Persons with DS-pulmonary TB were selected from a randomized, double blind controlled trial of adjunctive high-dose cholecalciferol (vitamin D_3_) for TB treatment conducted in the country of Georgia (clinicaltrials.gov identifier NCT00918086) (*14*). All persons included in this metabolomics sub-study were HIV-negative. Inclusion criteria for patients included age ≥ 18 years and newly diagnosed active TB disease, based on a positive AFB sputum smear and confirmed by positive sputum culture for *Mtb*. Baseline plasma samples for HRM were obtained from eligible subjects within 7 days of initiating therapy with conventional dosing of first-line anti-TB drugs (isoniazid, rifampicin, pyrazinamide and ethambutol) (*14*). Phenotypic DST was performed on *Mtb* isolates recovered from all persons with pulmonary TB using the absolute concentration method (*34*).

### Controls without active TB disease

Plasma from persons with and without LTBI was analyzed for cross-sectional comparison with pulmonary TB cases. Persons with LTBI were enrolled from the DeKalb County Board of Health in DeKalb County, GA, USA. All persons with LTBI had positive test results from at least two FDA-approved tests for LTBI (QFT, TSPOT.TB [TSPOT] and/or tuberculin skin test [TST]). All tests were interpreted according to the guidelines from the Centers for Disease Control and Prevention (*35 36*). Controls without LTBI were U.S.-born adults at low risk for *Mtb* exposure and infection, who had at least one negative TST documented in medical records (*37*).

### Plasma metabolomics analysis

De-identified samples were randomized by a computer-generated list into blocks of 40 samples prior to transfer to the analytical laboratory where personnel were blinded to clinical and demographic data. Thawed plasma (65 μL) was treated with 130 μl acetonitrile (2:1, v/v) containing an internal isotopic standard mixture (3.5 μL/sample), as previously described (*38*). The internal standard mix for quality control consisted of 14 stable isotopic chemicals covering a broad range of small molecules (*38*). Samples were mixed and placed on ice for 30 min prior to centrifugation to remove protein. The resulting supernatant was transferred to low-volume autosampler vials maintained at 4°C and analyzed in triplicate using an Orbitrap Fusion Mass Spectrometer (Thermo Scientific, San Jose, CA, USA) with c18 liquid chromatography (Higgins Analytical, Targa, Mountain View, CA, USA, 2.1 x 10 cm) with a formic acid/acetonitrile gradient. The high-resolution mass spectrometer was operated in negative electrospray ionization mode over scan range of 85 to 1275 mass/charge (*m/z*) and stored as .Raw files (*39*). Data were extracted and aligned using apLCMS (*40*) and xMSanalyzer (*41*) with each feature defined by specific *m/z* value, retention time and integrated ion intensity (*39*). Three technical replicates were performed for each plasma sample and intensity values were median summarized (*42*).

### Metabolite identification and reference standardization

Identities of metabolites of interest were confirmed using ion dissociation methods (tandem MS/MS). Fragmentation spectra were generated using a Q Exactive HF Hybrid Quadrupole-Orbitrap Mass Spectrometer (Thermo Fisher Scientific, Waltham, MA) with parallel reaction monitoring mode using a targeted inclusion list. TCA cycle metabolites and arachidonic acid were confirmed and quantified by accurate mass, MS/MS and retention time relative to authentic standards (17).

### Untargeted lipidomics

Lipids were extracted from each plasma sample using a high throughput methyl t butyl ether (MtBE) extraction procedure with an automated and robust liquid handling instrument (Biotage Extrahera, Uppsala, Sweden) as previously described (*43*). Extracted samples were dried under nitrogen and reconstituted in 200 μl 1:1 chloroform:methanol prior to injection into the LC/MS system and lipids were resolved using a Thermo Acclaim C18 reverse phase column on a Thermo Vanquish UPLC coupled to a Thermo Fusion IDX mass spectrometer (Thermo, Waltham, MA) (*44*). Data were acquired at full scan mode at a resolution of 240,000 FWHM for all the samples and iterative data dependent acquisition (DDA) mode was collected on pooled samples at a resolution of 35,000 step-wise collision energy for identification of lipids. Data was processed using LipidSearch (Thermo Fisher, San Jose, CA). Lipids that contained a signal to noise ratio of greater than 10 and had a high confidence level (MS/MS) identifications with CVs less than 30% across pooled QCs were considered for downstream analysis.

### Targeted measurement of eicosanoids

For targeted analysis of oxylipins, these lipids were enriched from bulk membrane lipids by performing solid phase extraction using the Biotage Extrahera liquid handling system as previously described (*23, 45, 46*). The resulting extracts were analyzed using a multiple reaction monitoring (MRM)-based LC/MS protocol on a QTrap 5500 (Sciex, Waltham, MA) synced to Exion LC AD system (SCIEX, Waltham, MA), whereby detected oxylipins and endocannabinoids were fragmented and quantified against external standard curves. Oxylipins and endocannabinoids detected in less than 60% of patient samples were excluded from this analysis. Missing values were imputed using half of the minimum detected value for each lipid.

### Plasma cytokine detection

The U-PLEX assay (Meso Scale MULTI-ARRAY Technology) commercially available by Meso Scale Discovery (MSD) was used for plasma cytokine detection. This technology allows the evaluation of multiplexed biomarkers by using custom made U-PLEX sandwich antibodies with a SULFO-TAG™ conjugated antibody and next generation of electrochemiluminescence (ECL) detection. The assay was performed according to the manufacturer’s instructions (https://www.mesoscale.com/en/technical_resources/technical_literature/techncal_notes_search). In summary, 25μL of plasma from each participant was combined with the biotinylated antibody plus the assigned linker and the SULFO-TAG™ conjugated detection antibody; in parallel a multi-analyte calibrator standard was prepared by doing 4-fold serial dilutions. Both samples and calibrators were mixed with the Read buffer and loaded in a 10-spot U-PLEX plate, which was read by the MESO QuickPlex SQ 120. The plasma cytokines values (pg/mL) were extrapolated from the standard curve of each specific analyte.

### Statistical analysis

Statistical comparisons of metabolite and lipid intensity values (abundance) and concentrations were performed in R version 3.6.1. For untargeted metabolomics and lipidomics analyses, metabolite intensity values were log_2_ transformed and compared between groups using linear regression, controlling for age, sex and HIV status (*47*). Metabolic pathway enrichment analysis was performed using *mummichog*, a Python-based informatics tool that leverages the organization of metabolic networks to predict functional changes in metabolic pathway activity (*15, 30, 48*). Following quantification of selected metabolites and lipids, cross-sectional comparison of plasma concentrations between groups was made using the Wilcoxon Rank Sum test. Changes relative to baseline during treatment of TB disease were tested using a Wilcoxon Signed-Rank test. For correlation analyses, plasma metabolite, lipid and cytokine concentrations were normalized using log transformation. A p-value less than or equal to 0.05 was considered statistically significant. For untargeted analyses, a false discovery rate (FDR) of q<0.05 was used (*19*).

### Study Approval

All studies were approved by the Institutional Review Board (IRB) of Emory University (Atlanta, GA, USA), and by the individual IRBs associated with the original cohort studies: the Ethics Committee of the National Center for Tuberculosis and Lung Diseases of Georgia (Tbilisi, Georgia), the University of KwaZulu-Natal IRB (Durban, South Africa), and the Georgia Department of Public Health IRB (Atlanta, GA, USA), respectively, depending on the site of participant enrollment. All subjects provided written informed consent.

## Data Availability

All study data will be made available by the authors upon reasonable request.

## SUPPLEMENTARY MATERIALS

**Fig. S1.**
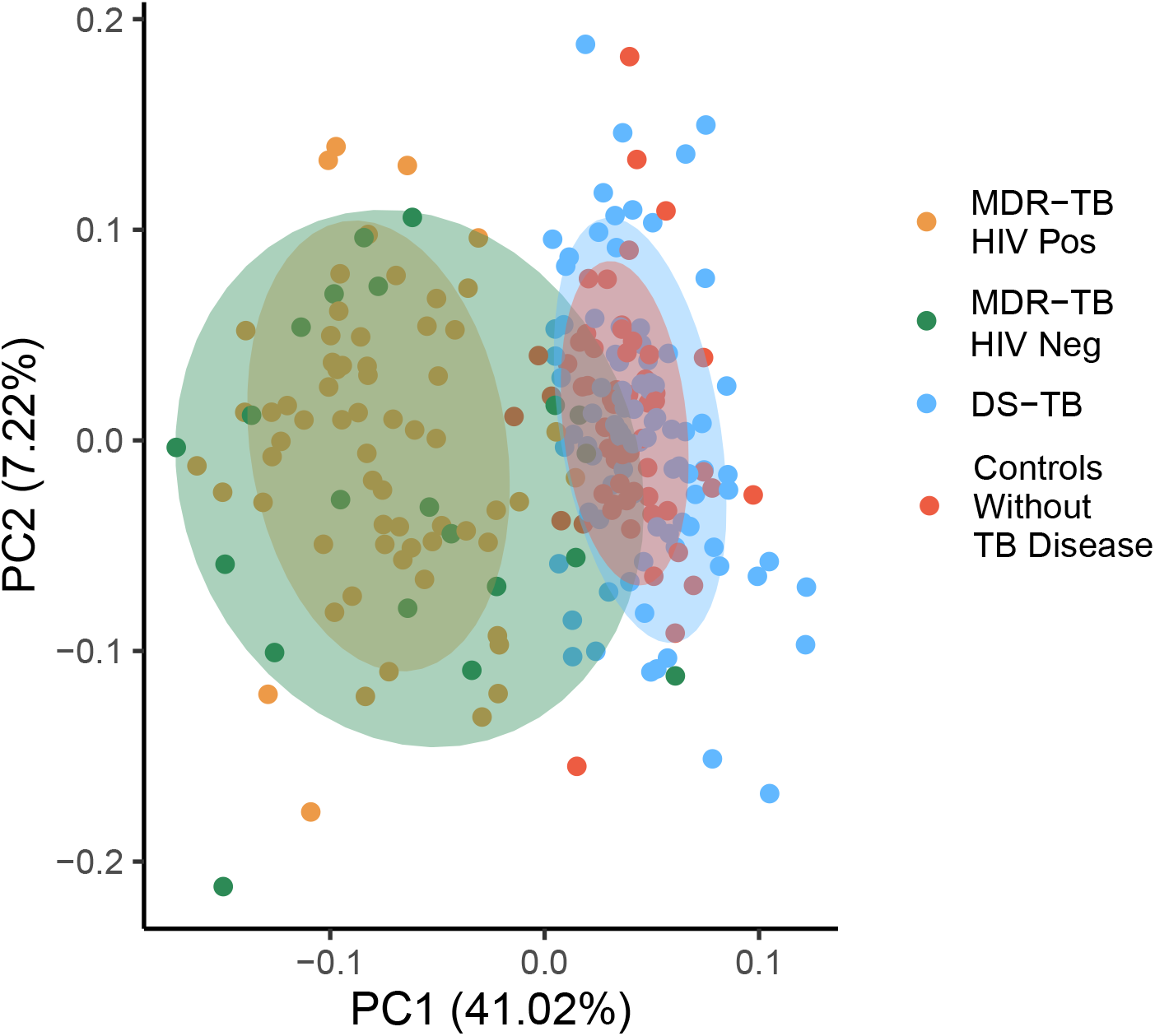
Effect of HIV co-infection on metabolic changes in MDR-TB. Principal component analysis of metabolites mapping to significantly regulated pathways in persons with multidrug resistant (MDR)-TB. Person with MDR-TB with (n=64; yellow) and without (n=21; green) HIV co-infection clustered together and both groups clearly separated from persons with drug-susceptible (DS)-TB (n=89; light blue) and controls without TB disease (n=57; red).

**Fig. S2.**
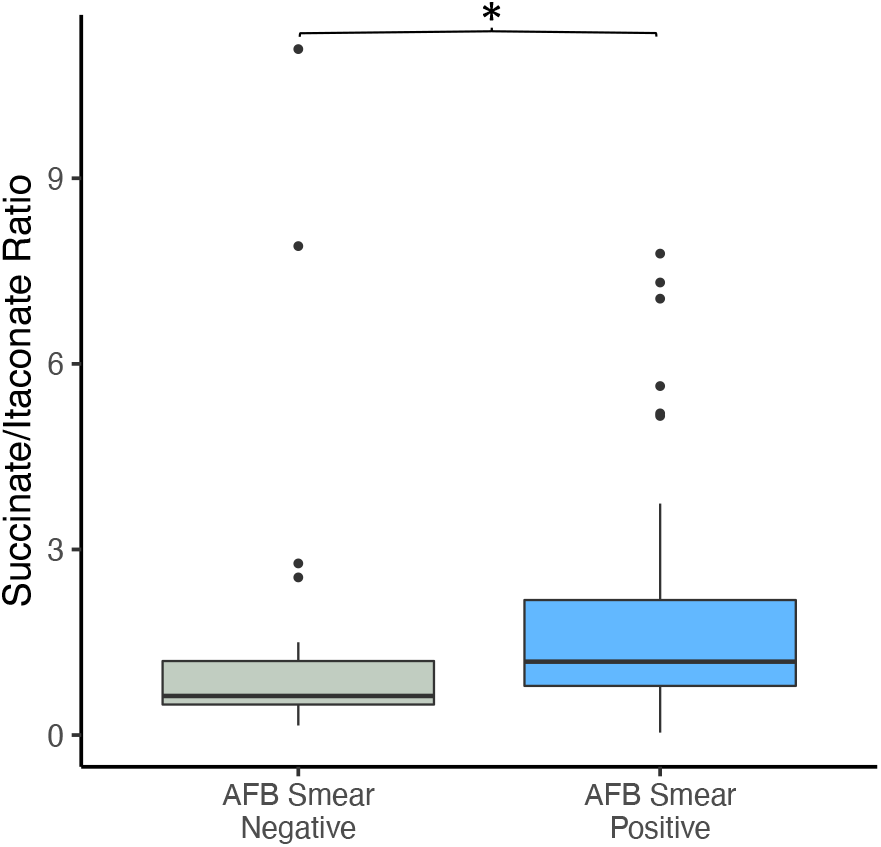
Plasma ratio of succinate to itaconate in drug susceptible TB. In persons with drug susceptible (DS)-TB, the plasma ratio of succinate to itaconate was significantly increased in those with a persistently positive sputum smear for acid-fast bacilli (AFB) at enrollment versus those who converted to a negative AFB sputum smear.

## Acknowledgements

We thank all the study teams and study sites from the multiple cohorts leveraged by this metabolomics study, including the Georgia National Center for Tuberculosis and Lung Disease, the University of KwaZulu-Natal and the DeKalb County Board of Health, Georgia, USA.

## Funding

National Institutes of Health grant R21 AI130918 (TRZ, NRG, JCMB, DPJ, RRK, HMB) National Institutes of Health grant K23 AI144040 (JMC)

National Institutes of Health grant P30 AI050409 (JMC),

National Institutes of Health grant T32 AI074492 (JMC),

National Institutes of Health grant P30 ES019776 (DPJ, TRZ)

National Institutes of Health grant R01 AI087465 (NRG, NSS, JCMB)

National Institutes of Health grant K24 AI114444 (NRG)

National Institutes of Health grant D43 TW007124 (RRK, HMB)

National Institutes of Health grant R01AI114304 (JCMB)

National Institutes of Health grant R01AI145679 (JCMB)

National Institutes of Health grant K24AI155045 (JCMB)

National Institutes of Health grant P30AI124414 (JCMB)

National Institutes of Health grant UL1 TR002378 (JMC, MK, HMB, EAO, TRZ)

National Institutes of Health grant UL1TR001073 (JCMB)

Emory University Global Health Institute (TRZ, HMB, NT, RRK)

Emory Medical Care Foundation (RRK, JMC, HMB, TRZ)

## Author contributions

Conceptualization: JMC, DPJ, JCMB, RPS, NRG, HMB, EAO, TRZ

Methodology: JMC, DPJ, AS, MK, KHL, KM, RPS, EAO, TRZ

Investigation: JMC, AS, MK, KHL, KM, BP,

Visualization: JMC, AS, KHL, BP, RPS, TRZ

Funding acquisition: JMC, DPJ, MK, RRK, NT, JCMB, NRG, HMB, EAO, TRZ

Project administration: JMC, DPJ, RRK, RPS, HMB, EAO, TRZ

Supervision: DPJ, RRK, JCMB, RPS, NRG, HMB, EAO, TRZ

Writing – original draft: JMC

Writing – review & editing: JMC, DPJ, AS, MK, RRK, BP, KM, NSS, JCMB, RPS, NRG, HMB, EAO, TRZ

## Competing interests

Authors declare they have no competing interests.

## Data and materials availability

All data, code and materials from this analysis will be made available by the authors upon reasonable request.

## Notes

### Competing Interest Statement

The authors have declared no competing interest.

### Funding Statement

This work was supported in part by grants from the NIH including R21 AI130918 (T.R.Z., N.R.G., J.C.M.B., D.P.J., R.R.K., H.M.B.), R01 AI087465 (N.R.G, N.S.S., J.C.M.B.), T32 AI074492 (J.M.C.), K23 AI144040 (J.M.C.), K24 AI114444 (N.R.G.), Georgia Clinical and Translational Science Alliance UL1 TR002378 (T.R.Z., H.M.B., J.M.C.), as well as grants from the Emory University Global Health Institute (T.R.Z., H.M.B., N.T., R.R.K.), and the Emory Medical Care Foundation (R.R.K., J.M.C., H.M.B., T.R.Z.).

